# Heat-based Decontamination of N95 Masks Using a Commercial Laundry Dryer

**DOI:** 10.1101/2020.07.22.20160283

**Authors:** Yuri D. Lensky, Edward A. Mazenc, Daniel Ranard, Matthew Vilim, Manu Prakash, Bill Brooks, Amanda Bradley, Alain Engelschenschilt, Jason Plutz, Todd Zellmer

**Affiliations:** Stanford University, Stanford CA 94305 USA; Alliance Laundry Systems, Ripon WI 54971 USA

## Abstract

We propose a dry heat method for decontaminating N95 masks of SARS-CoV-2, designed around placing them in resealable plastic bags, packed in large cardboard boxes installed at the rear end of commercial laundry dryers. Our protocol rests on data collected in collaboration with Alliance Laundry Systems (ALS) and the CDC/NIOSH laboratories, under the “NPPTL Respirator Assessments to Support the COVID-19 Response” initiative. We test the two most widely available ALS tumbler models, the UTF75N and UT075N, and show that if our procedure is carefully followed, the masks will be subject to suitably high and stable temperatures for decontamination; in particular the masks will be heated to at least 80 °C for at least 65 min. For the mask models 3M 1860, 3M 8511, and Halyard 62126, we establish that they pass quantitative fit tests and retain sufficient filtration performance after three cycles of our decontamination procedure. All masks used in this study were new and uncontaminated: the evidence for the levels of biological inactivation of SARS-CoV-2 is provided by [1]. While the protocol outlined here is currently specific to certain tested dryer models, this equipment is widely available, with machines estimated to be within 15 minutes of most US hospitals. Models from other manufacturers may also be appropriate for this decontamination method, though we stress the need for explicit testing on alternative models before use.

## 1 DISCLAIMER

The Content provided here is for INFORMATIONAL PURPOSES ONLY and DOES NOT CONSTITUTE THE PROVIDING OF MEDICAL ADVICE and IS NOT INTENDED TO BE A SUBSTITUTE FOR INDEPENDENT PROFESSIONAL MEDICAL JUDGMENT, ADVICE, DIAGNOSIS, OR TREATMENT. Use or reliance on any Content provided here is SOLELY AT YOUR OWN RISK.

## 2 Overview & Summary of Results

The COVID-19 pandemic has led to mass shortages of personal protective equipment (PPE), most notably N95 filtering facepiece respirators (FFRs). When new N95 FFRs are unavailable, health care practitioners see themselves forced to reuse PPE potentially contaminated with SARS-CoV-2, the viral strain leading to COVID-19 disease, as well as other pathogens.

There are multiple decontamination methods available for the treatment of N95 FFRs. These decontamination methods include Hydrogen Peroxide Vapor (HPV), irradiation by UV-C light, and heat, amongst others (for reviews of literature on all methods see [2, 3]). Several hospitals with access to HPV treatment have adopted this method, which reduces infective viral loads by at least six orders of magnitude while preserving FFR performance. In our view, the promise of dry heat based methods lies in their global scalability, as well as relative simplicity and robustness of implementation, especially in settings where more sophisticated equipment may not be readily available. These might include smaller remote hospitals, skilled nursing facilities, correctional institutions, and cruise ships, as well as communities worldwide with lower resources.

The main purpose of this paper is to propose a dry-heat decontamination method, using industrial dryers as the heat source. The biological basis of our method is the work of Fisher et. al [1], who studied the effects of 60 min dry heat at 70 °C for the inactivation of SARS-CoV-2 directly on N95 FFRs. They report a greater than 3 log_10_-reduction in the viricidal activity of SARS-CoV-2 on the mask. We stress our protocol relies entirely upon this result to gauge its decontamination ability. We chose the parameters of our method so that taking the worst-case measurements (as discussed in more detail in Section 4.1), masks are subject to temperatures greater than 80 °C for more than 65 min. Even in the worst case, there are safety margins for error of 10 °C and 5 min above the Fisher result.

The bulk of the paper is organized into two main parts. First, we outline a specific protocol, in enough detail to allow on-site agents to design an appropriate Standard Operating Procedure (SOP). Second, we present the collected data which informed the design of this protocol. The experimental results of this paper fall into two broad categories: 1) the heat distribution and stability in specific models of commercial laundry dryers and 2) the effects of the decontamination procedure on the filtration, fit and elastomeric integrity of the FFRs after 3 decontamination cycles.

The first data set was collected primarily in the research and development labs of Alliance Laundry Systems, as well as one field installation. We test their two most widely available tumbler models (i.e. tumble laundry dryers), the UTF75N and UT075N. We present these results in section 4.1. We highlight potential pitfalls of the method, such as the presence of significant temperature gradients within the dryer. We discuss how we addressed such pitfalls in the design of our protocol. The apparent ease of using heat as a decontamination method can be misleading: one must guarantee stable and homogeneous temperatures within a prescribed temperature range for the safe processing of PPE. Because the protocol below is model-specific, we hope other groups perform similar assessments of this method for dryers by different manufacturers.

The second data set focuses on the effects of the decontamination procedure on the FFR’s fit and filtration efficiency. It was provided to us by Michael Bergman’s group at the National Personal Protective Technology Laboratory (NPPTL) under the special initiative “Assessment of filter penetration performance and fit for decontaminated N95 respirators” [5]. They tested 20 samples of three models of FFRs (3M 1860, 3M 8511 & Halyard/Kimberly-Clark 62126) which had undergone three cycles of our decontamination procedure. We summarize their findings, as well as their experimental setup and methodology, in section 4.2. Most importantly, all three models had at least 95 % filter efficiency and passing fit tests after 3 cycles. The quantitative filtration results are summarized in Table 2, and the raw data of the full assessment can be found in Appendix B. We hope to include results on higher cycle numbers in the near future.

We briefly comment on the availability of the tested dryer models as pertaining to the viability of this method, before concluding in section 5. We close by emphasizing once more that this decontamination proposal should in principle work on models from manufacturers other than ALS, but that testing needs to be performed on each model before usage to ensure safe processing of PPE.

## 3 Protocol

### Cautionary Note

*Laundry dryers are not designed for medical decontamination procedures. Most models will likely suffer from large temperature gradients across the tumbler and not reach sufficiently elevated temperatures. This data-driven protocol should therefore not be used on dryer models not explicitly addressed in Appendix A, nor modi.fled in any way from its current form, until it has been tested for safe usage on other machines*.

Based on the experiments and existing data we discuss in Section 4, we propose the following protocol for mask decontamination. This protocol is provided as a skeleton for a more detailed Standard Operating Procedure (SOP) to be devised on-site, so as to best accommodate local conditions. This protocol was designed for masks without visible soiling; larger amounts of contaminated substrate may have a protective effect on SARS-CoV-2 and reduce the efficacy of the heat-based decontamination [3]. Moreover, we re-emphasize that this protocol is only designed for the inactivation of SARS-CoV-2 rather than other pathogens.

### 3.1 Required Equipment

- 3-4 bungee cords
  – Hooks must fit through the tumbler cylinder grating.
  – Cords should be taut when stretched to cylinder diameter.
- “Ziploc Grip-’n-Seal” 1 gallon freezer bags^1^
- Black Gorilla Tape^®^
- Compatible commercial laundry dryer. To determine if a model is compatible, as well as the necessary temperature and time settings, see Appendix A. This method can only be used with models explicitly addressed in Appendix A.
- 2 single wall corrugated cardboard boxes of dimensions 16 × 16 × 16 in. Our tests were done with Uline S-4166 boxes, and we recommend these exact boxes are used. We believe, but have not yet experimentally confirmed, that other single wall corrugated cardboard boxes of the same dimension produce similar results. A diagram of a single box is shown in Figure 1.

**Figure 1:**
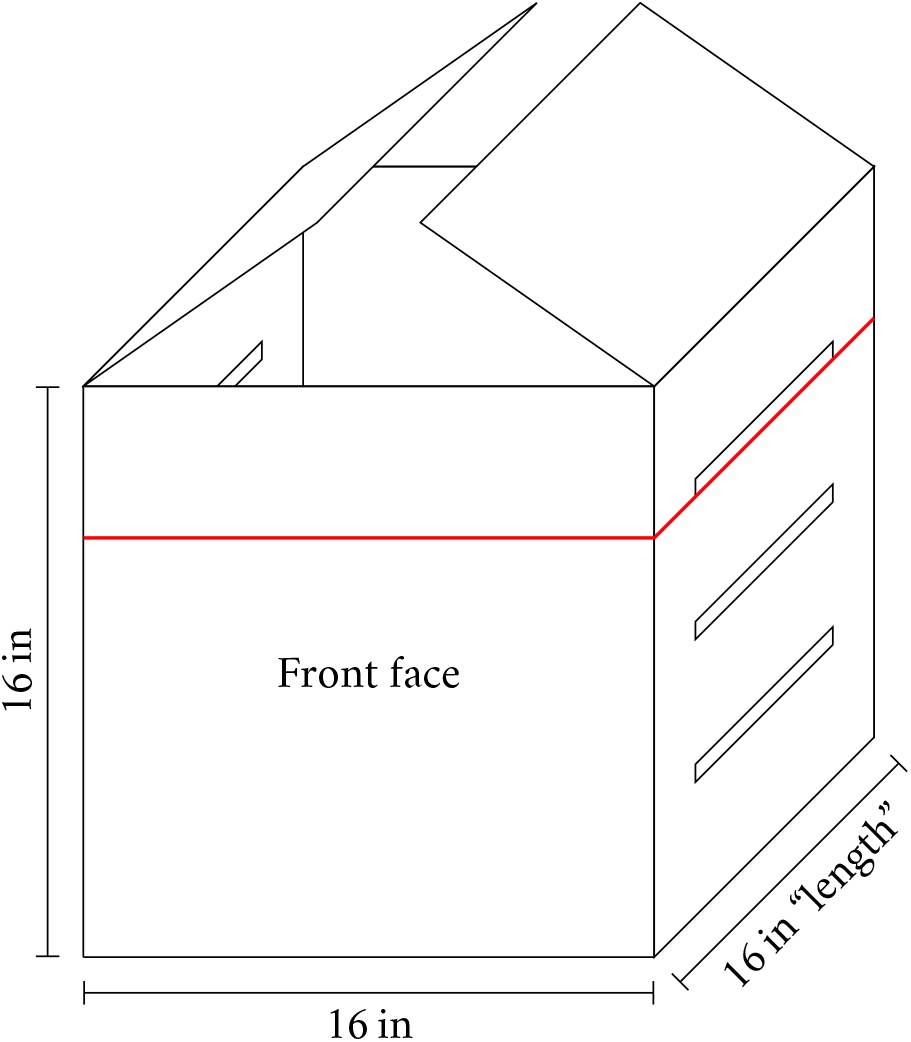
Labeled diagram of a single cardboard box used in the protocol. The three 10 × 0.5 in slits to be cut by the user are shown on the side face (note that there are another 3 on the opposite face, for a total of 6). They should be evenly spaced, with centers 4 in, 8 in, and 12 in from the bottom of the box. The maximum height to which the bagged masks can fill the box is shown by a red line.

### 3.2 Protocol Steps

1. For each N95 FFR, check that it has not been decontaminated more than twice. Since masks can only be decontaminated a limited number of times, it is important to track how many times they have been run through a decontamination cycle. A simple method is to use a permanent marker on the mask to tally the number of decontamination cycles. If the mask can be decontaminated, mark or otherwise update the tracking of the mask at this step (for example, if a mask had 2 tallies before this step, it should have 3 after this step).
2. On each of the 2 side faces of the box, cut 3 slits of dimension 10 × 0.5 in (6 slits in total). The slits should be evenly spaced, with centers 4 in, 8 in, and 12 in from the bottom of the box. See Figure 1.
3. Place used N95 FFR in Ziploc bag and seal said bag. New bags should be used for each round of decontamination. Information identifying the user of the mask should be written on the bag, or written on the mask with permanent marker. If the model includes a metallic nose piece, wipe the nose piece with a disinfectant wipe.^2^ See Fig.2a.
4. Place the Ziplocs containing masks inside the cut cardboard boxes. The number of masks should be small enough that when resting on the ground, the masks do not fill the box past the topmost slot. In any case, there should be no more than 20 masks per box. Tape the boxes shut with Black Gorilla Tape^®^. See Figures 1 and 2b.
5. For this step, we refer to Figures 1 and 2c. Place the first box against the rear of the tumbler, so that the “Front face” faces out of the tumbler, and the box is in the orientation shown in Figure 1. Place the second box on top of the first, so that the slits face out to the sides.
6. Affix the bungee cords to create a barrier; see Figure 2c. The bungee cord barrier should be located (16.5 to 16.75) in from the rear.
7. Set temperature and time settings according to the model information in Appendix A. In particular, all tested models have required main cycle time setting 99 min.

## Evidence for viability of protocol

### 4.1 Characterization of dryer temperatures

As discussed in Section 4.3, SARS-CoV-2 is inactivated when subjected to high enough temperatures for sufficient time. On the other hand, the performance of N95 FFRs can degrade under high enough heat [6], so an ideal heat-based decontamination method must stably maintain temperatures in a narrow window. In this section, we describe experiments demonstrating that under suitable conditions, certain industrial dryers can meet these requirements. The experiments were performed at an ALS testing facility on two machines, with tumbler model numbers UT075N and UTF75N, as well as a standard UTF75N tumbler installation at a hotel. We will also refer to these as the “normal” (UT075N) and “fast” (UTF75N) tumblers. Both tumblers have 75 pound capacity and a natural gas heating element, but the “fast” model has a more powerful heating element, rated for 65.9 kW fuel input, compared to the “normal” 48.3 kW.

**Figure 2:**
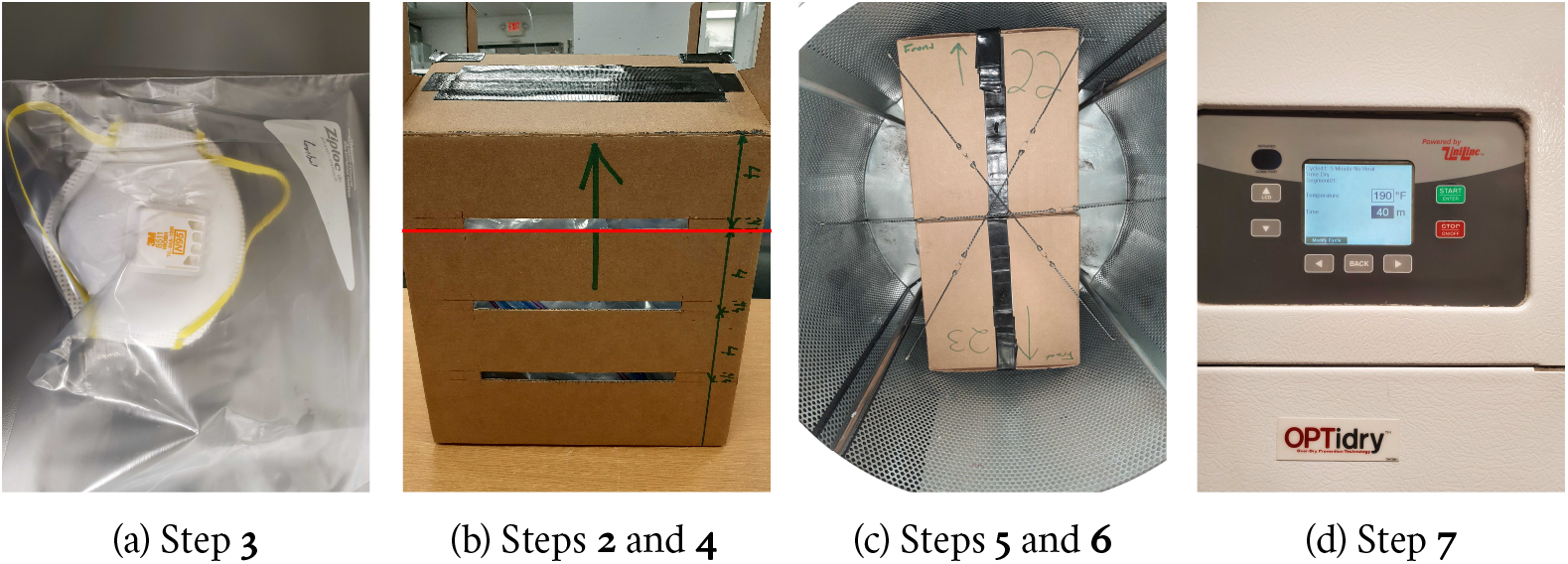
Illustrations of steps in the protocol. In Figure 2b, we show the maximum filling height by a red line (see also Figure 1).

Dryers are not designed to maintain a narrow temperature range, or to be run without damp linens. To characterize the typical behaviour of the dryer under these unusual conditions, we mounted 9 HOBO^®^ U12-015 temperature loggers along the dryer cylinder as shown in Figure 3. These loggers have stated accuracy ±0.75 °C below 100 °C, and ±0.65 °C below 90 °C^3^. The mean temperatures are shown in Figure 4. The most extreme temperature gradients are more than 45 °C over 10 in (more than 50 °C across the tumbler), which is well beyond the acceptable limit for a heat decontamination protocol.

**Figure 3:**
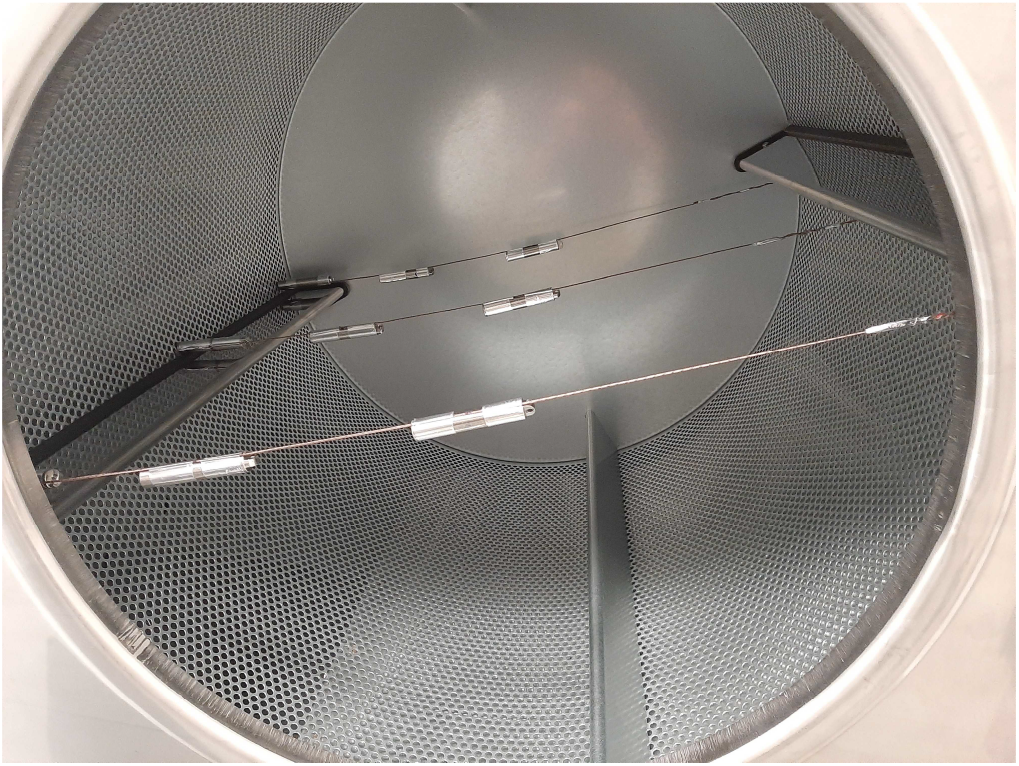
Installation of temperature sensors throughout dryer tumbler. 3 sensors installed in front, middle and rear sections, radially distributed to probe temperature variations from central axis to dryer wall.

**Figure 4:**
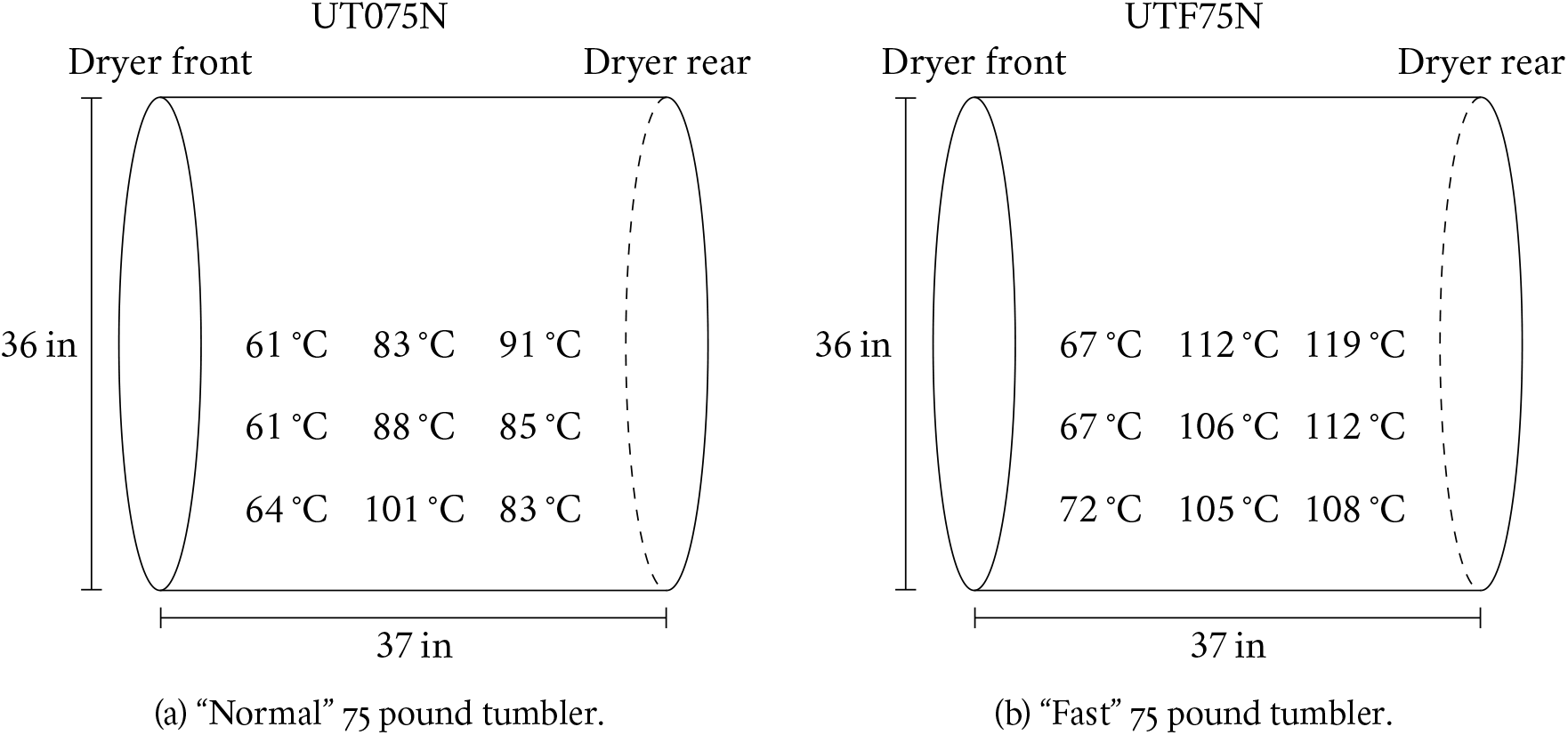
Radial mean temperature distribution in the dryer cylinder at the 190 °F (88 °C) setting.

Our protocol is designed around establishing a region of appropriate temperature within the dryer. In particular, we place the masks in cardboard boxes and restrict the boxes to the rear half of the dryer. The basic idea is to keep the masks protected from the extreme temperatures on the cylinder itself, and to stabilize the temperature both by using the region in the tumbler with lower temperature gradients and by using the more uniform equilibrium in the cardboard box. An added benefit of using cardboard boxes is to reduce impact on the masks from the tumble.

To test these ideas directly, we mounted 2 HOBO sensors in 2 cardboard boxes, as shown in Figure 5. We then follow the steps of our protocol outlined in Section 3.2, except for two important differences. First, we partially fill the Ziploc bags with air instead of masks (see Figure 5c). For users testing other models, we point out that the presence of the Ziplocs is critical to get an accurate characterization of the time to reach a given temperature, although they do not have a significant effect on the equilibrium temperature (data not shown).

**Figure 5:**
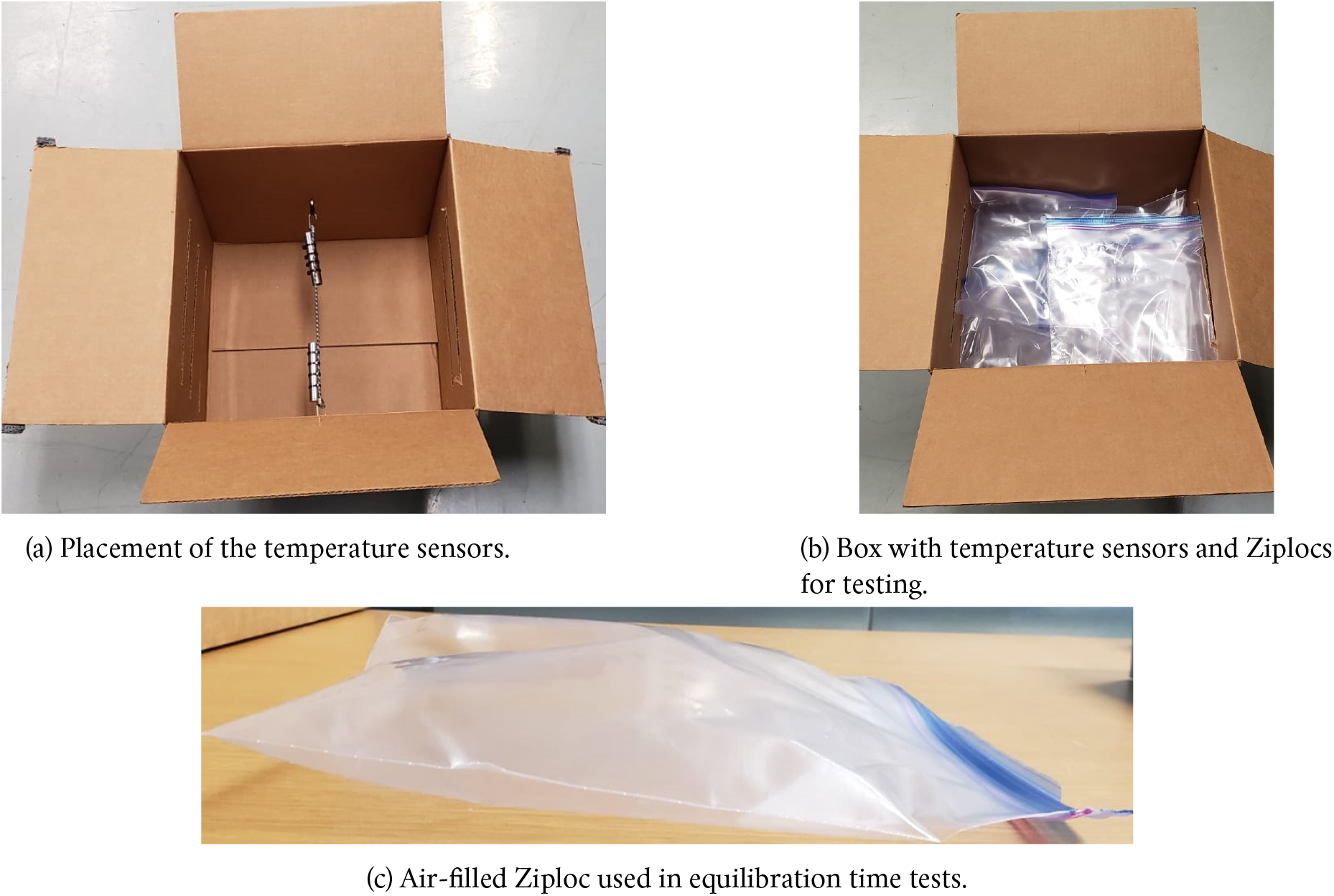
Experimental setup for temperature readings used to test protocol.

Second, our protocol dictates runtimes of 99 min, but most of our testing is done at runtimes of 60 min. We found that during each test, for each HOBO sensor there is a time, called the “equilibration time,” and a temperature, called the “equilibrium temperature,” such that all temperatures measured on that sensor after the equilibration time are within the HOBO accuracy of 0.65 °C of the equilibrium temperature. Among all of our measurements (both 60 min and 99 min on all machines), the maximum equilibration time is 53 min. Thus we performed a single test at 99 min to check that equilibrium temperatures do not drift or fluctuate over the course of a longer run (equivalently, that the equilibration time for a 99 min run is less than 60 min). We then extrapolate results from our 60 min runs to 99 min at the accuracy of the HOBO sensor simply by assuming the sensors will maintain their equilibrium temperatures. We note that, since the minimum equilibrium temperature measured over all runs is greater than 83.15 °C and the longest time to 80 °C is 30.5 min, even with the weaker assumption that there are no fluctuations greater than 2.5 °C for more than 3.5 min after equilibration time (see Table 1) our protocol still achieves at least 80 °C for more than 65 min in the worst-case conditions we measured.

**Table 1:** Summary of protocol properties. We show both the raw values, and values conservatively corrected for our sensor equilibration time (as discussed in footnote 4). For the UTF75N tumbler type, the data are reported over 7 separate runs at a 165 °F main cycle setting, with a total of 2 60 min runs (8 sensor logs) in a hotel installation, 4 60 min runs (16 sensor logs) in the ALS laboratory installation, and 1 99 min run (3 sensor logs due to a sensor failure) in the ALS laboratory installation. For the UT075N tumbler type, the data are reported over 3 separate runs in an ALS laboratory installation at a 170 °F, with 2 60 min runs (8 sensor logs) and 1 99 min run (4 sensor logs). The time to 80 °C is the time after which the temperature never falls below 80 °C; the raw maximum time is taken over all HOBO sensor logs from the relevant tumbler and rounded up (to the nearest integer minute above). The contribution of systematic errors from the sensors (in both time and temperature) are below our precision for all reported values. The estimated mean (standard deviation) of the distribution underlying a quantity is shown before (following) the ± symbol. The standard deviation dominates the uncertainty in each estimated mean, which we show in parentheses following the estimate. The fluctuations about the equilibrium temperature of a given sensor are below systematic errors of the HOBO sensors, but the raw equilibration times are below 60 min (maximum over all runs and sensors is 53 min), which justifies our use of 60 min runs to evaluate the behaviour over 99 min (see main text for definitions of equilibration time and equilibrium temperature and further discussion). The mean equilibrium temperature is defined as the mean of the equilibrium temperatures of all HOBOs in a given experimental run, and likewise for the mean equilibration time. Importantly, we find that the hotel installation of the UTF75N tumbler behaves similarly to (i.e. within statistical fluctuations and measurement uncertainty of) the ALS installation.

| Tumbler # | Max time to 80 °C (raw/corrected min) | Min projected time at 80 °C for a 99 min run (raw/corrected min) | Time to 80 °C (raw/corrected min) | Mean equilibrium temperature (°C) | Mean equilibration time (raw min) |
| --- | --- | --- | --- | --- | --- |
| UTF75N (hotel) | 31/27.5 | 68/71.5 | 26.8/23.3(1.1) ±3.0 | 84.7(1.4) ±1.9 | 38.1(3.1) ±4.4 |
| UTF75N (ALS) | 33/29.5 | 66/69.5 | 26.3/22.8(0.9) ±3.9 | 86.4(0.3) ±0.7 | 42.6(2.0) ±4.6 |
| UTF75N (all) | 33/29.5 | 66/69.5 | 26.5/23.0(0.7) ±3.6 | 85.9(0.5) ±1.3 | 41.3(1.8) ±4.7 |
| UT075N | 34/30.5 | 65/68.5 | 27.9/24.4(0.9) ±3.2 | 85.4(0.2) ±0.3 | 43.0(1.9) ±3.3 |

Our final protocol was tested (with the modifications described above) 7 times in the case of the UTF75N model, corresponding to 27 sensor logs (4 sensors per run with 1 sensor failure), and 3 times in the case of the UT075N model, corresponding to 12 sensor logs. The 99 min runs for the two types of tumbler (UTF75N and UT075N), as well as sample 60 min runs, are shown in Figure 6. Our key findings Placement of the temperature sensors. (b) Box with temperature sensors and Ziplocs for testing. are summarized in Table 1. The most important figure is the worst-case time, measured over all experiments, to reach 80 °C. We show this time both raw, and corrected for the HOBO sensor equilibration time of 3.5 min^4^. This figure provides an accurate under-estimate of the time the masks will spend at or above 80 °C; we have chosen our temperature settings so that the HOBO sensors never dip below 80 °C throughout the run after reaching it. This demonstrates the viability of our method; we achieve more than 80 °C for more than 65 min even in the worst case. Finally, the agreement in Table 1 between the hotel and ALS UTF75N tumblers provides evidence that our method produces similar results across proper installations of the same tumbler type.

**Figure 6:**
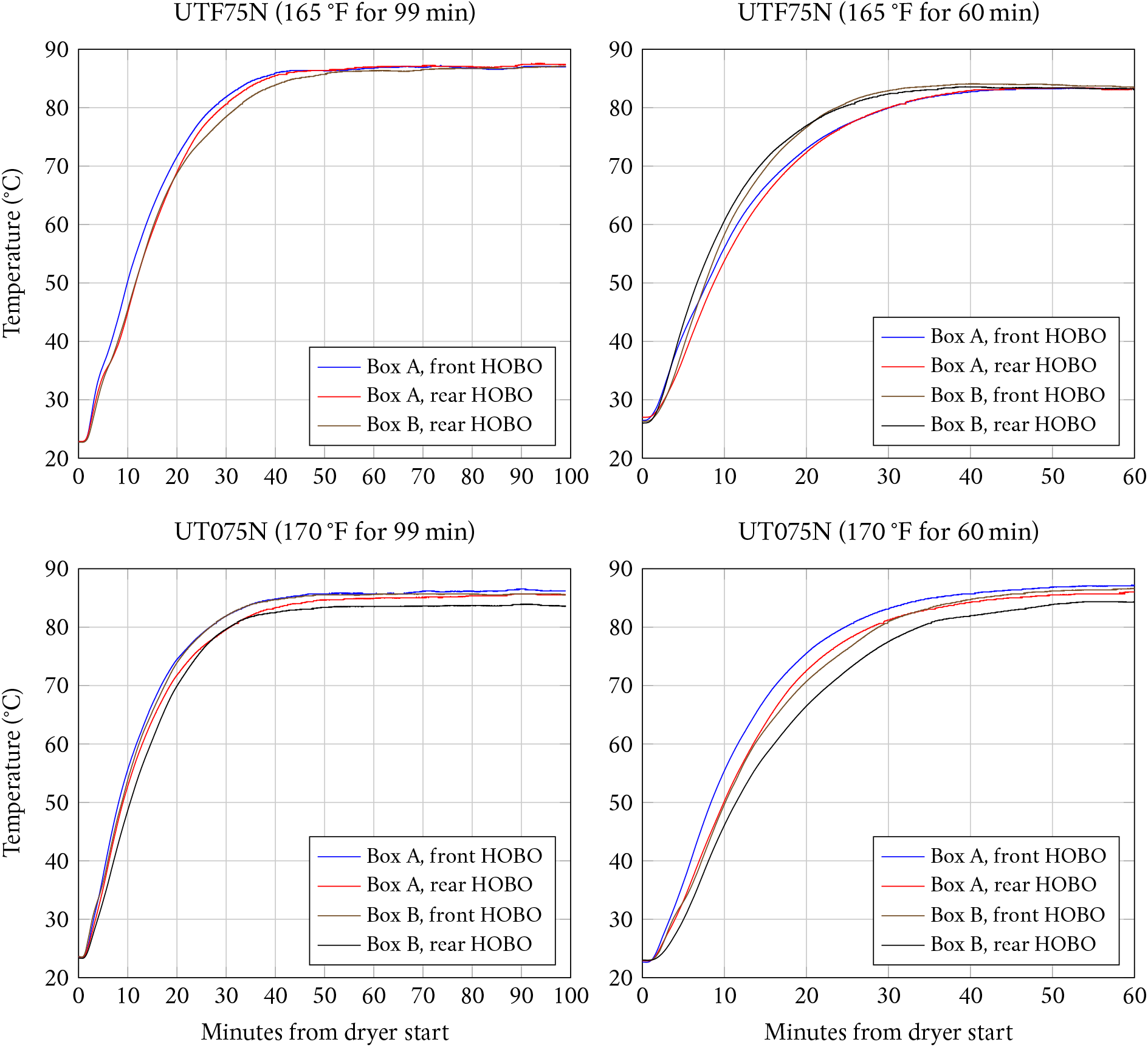
Data from the monitored dryer runs, labeled by the tumbler model number, and main cycle temperature and time settings. Note that one HOBO sensor failed in the 99 min UTF75N run.

### 4.2 Mask integrity

In addition to potential damage to N95 FFRs from heat treatments [6], one may also be concerned that other aspects of our decontamination procedure, such as the impact from tumbling or rubbing against Ziploc bags, have detrimental effects on mask performance. To assess the potential for damage, we performed 3 cycles of our treatment on 15 samples of 3 types of masks (45 treated masks in total). The three models are 1. 3M 1860, 2. 3M 8511, and 3. Halyard 62126, which were run under appropriate settings in a UTF75N machine to subject them to at least 80 °C for at least 65 min.

We note that there were some differences in the procedure we tested on the masks to the one we characterize in Sections 3 and 4.1. The boxes we used were 16 × 14 × 17 in, and had only 2 slits. In this configuration, the temperature setting had to be made 170 °F (as compared to our new setting 165 °F) in order to ensure temperatures at least 80 °C for at least 65 min as equilibration time was slower on average, as well as less stable. The higher temperature setting meant that the equilibrium temperatures were significantly higher, of order ∼ 90 (2) °C. We prefer our improved method since it more reliably reaches 80 °C in short enough time, at lower equilibrium temperatures, as well as enabling use of the boxes for more runs due to reduced tumble impact on the boxes at our prescribed size. Importantly, both the impact of tumbling on the boxes and the equilibrium temperature are reduced in our new method, so mask integrity is preserved at least as well by our new method as the method used for the tests in this section.

The treated masks were then sent (along with 5 untreated control masks of each type) to the National Personal Protective Technology Laboratory (NPPTL), a research center within the National Institute for Occupational Safety and Health, to be tested under the special initiative “Assessment of filter penetration performance and fit for decontaminated N95 respirators” [5].

The NPPTL tests consist of filtration, fit, and tensile testing of the elastomeric straps. For each respirator type, 10 are used for filtration testing, 5 are used for fit, and 3 of the masks tested for filtration are also used for strap integrity testing. Five additional masks are used as controls. We emphasize that at no point was any data obtained for the same mask both before and after the treatment. Controls refer to masks of the same model which never underwent any decontamination heating. A brief summary of the key results was provided by the NPPTL, which we reproduce here:

#### Filtration efficiency

The minimum and mean filter efficiencies of the treated masks were ^5^:

**3M 1860** 95.08 % (minimum) and 98.05(0.40)% (mean)

**3M 8511** 98.14 % (minimum) and 98.78(0.10)% (mean)

**Halyard 62126** 97.31 % (minimum) and 98.41(0.19)% (mean).

**Manikin fit factor** The manikin fit factor showed passing fit factors (greater than 100) for all respirators evaluated. The manikin fit test procedure used in this assessment did not show any detriments in fit associated with the decontamination method used.

**Strap integrity** No visual degradation of the straps was observed. NPPTL reported the following relative decreases in recorded force for the top and bottom straps, respectively;

**3M 1860** 6.55 % and 2.53 %

**3M 8511** 12.09 % and −9.73 %; as noted by NPPTL, only in this mask type the results are inconsistent between the top and bottom straps, with the bottom straps getting *stronger*

**Halyard 62126** 4.61 % and 5.25 %.

While the exact correlation between the force exerted by straps and fit is not well understood, higher force values may be associated with a tighter fit of the respirator to the face.

In the following sections, we provide a brief overview of the NPPTL procedure for each type of test, and some comments on our results. The authoritative source on the NPPTL tests and relevant parameters is their test plan [5]. The complete raw data sent by NPPTL is displayed in Appendix B.

#### 4.2.1 Filtration efficiency

Particle filtration testing was run on 10 decontaminated masks of each type, as well as 3 control masks. The test is done on the TSI, Inc. automated filter tester, model 8130, which uses sodium chloride aerosol at mean particle size 0.075(0.020)µm, with flow rate set to 85.0(4.0)L min^−1^. The procedure mimics some parameters of the NIOSH Standard Test Procedure TEB-APR-STP-0059 [7], used to certify N95 FFRs, but the tests are not considered equivalent. For more details see [5]. The data for each mask type are summarized in Tables 4, 5, and 6. We summarize the most relevant data in Table 2.

**Table 2:** Summary of NPPTL filtration testing. The maximum efficiency drop is the difference between the most efficient control and least efficient treated mask. The mean efficiency drop is estimated as the difference of the control mean and mean for treated masks, and is shown to the left of the ± symbol. We estimate the standard deviation of the efficiency drop distribution as the square root of the sum of the variances of the treated and control data sets; assuming that the mask efficiency before and after decontamination is not anti-correlated, this is an overestimate of the true standard deviation. The estimated uncertainty in the mean efficiency drop is shown in parentheses, while the estimated standard deviation is shown following the ± symbol.

| Mask Type | Min efficiency (%) | Max efficiency drop (%) | Efficiency drop (%) |
| --- | --- | --- | --- |
| 3M 1860 | 95.08 | 4.05 | $0.95(0.40) \pm 1.26$ |
| 3M 8511 | 98.14 | 1.32 | $0.30(0.23) \pm 0.48$ |
| Halyard 62126 | 97.31 | 2.34 | $0.80(0.29) \pm 0.72$ |

**Table 3:** Decontamination settings for models whose temperature distribution was directly characterized as part of this work.

| Tumbler model # | Reversing | Main settings |  | Cooldown settings |  |
| --- | --- | --- | --- | --- | --- |
|  |  | Temperature (°F) | Time (min) | Temperature (°F) | Time (min) |
| UTF75N | No | 165 °F | 99 min | 70 °F | 5 min |
| UT075N | No | 170 °F | 99 min | 70 °F | 5 min |

**Table 4:** 3M 1860 filter efficiency evaluation.

| Sample # | Flow Rate<br>(l min<sup>-1</sup>) | Initial Filter<br>Resistance<br>(mmH<sub>2</sub>O) | Initial<br>Leakage (%) | Max Leakage<br>(%) | Efficiency (%) |
| --- | --- | --- | --- | --- | --- |
| 1 | 85 | 8.7 | 0.425 | 0.938 | 99.06 |
| 2 | 85 | 7.7 | 0.619 | 1.250 | 98.75 |
| 3 | 85 | 7.6 | 2.730 | 2.880 | 97.12 |
| 4 | 85 | 8.9 | 0.649 | 2.000 | 98.00 |
| 5 | 85 | 8.0 | 2.700 | 2.700 | 97.30 |
| 6 | 85 | 7.9 | 0.537 | 1.110 | 98.89 |
| 7 | 85 | 8.1 | 0.579 | 1.130 | 98.87 |
| 8 | 85 | 8.9 | 4.910 | 4.920 | 95.08 |
| 9 | 85 | 8.3 | 0.692 | 1.430 | 98.57 |
| 10 | 85 | 8.3 | 0.578 | 1.110 | 98.89 |
| Control 1 | 85 | 8.6 | 0.531 | 1.040 | 98.96 |
| Control 2 | 85 | 8.5 | 0.466 | 1.080 | 98.92 |
| Control 3 | 85 | 8.8 | 0.424 | 0.874 | 99.13 |

**Table 5:** 3M 8511 filter efficiency evaluation.

| Sample # | Flow Rate<br>(l min<sup>-1</sup>) | Initial Filter<br>Resistance<br>(mmH<sub>2</sub>O) | Initial<br>Leakage (%) | Max Leakage<br>(%) | Efficiency (%) |
| --- | --- | --- | --- | --- | --- |
| 1 | 85 | 7.6 | 0.355 | 1.070 | 98.93 |
| 2 | 85 | 6.9 | 0.496 | 1.350 | 98.65 |
| 3 | 85 | 6.9 | 0.577 | 1.160 | 98.84 |
| 4 | 85 | 6.1 | 0.348 | 1.060 | 98.94 |
| 5 | 85 | 7.6 | 0.385 | 1.040 | 98.96 |
| 6 | 85 | 7.4 | 1.370 | 1.860 | 98.14 |
| 7 | 85 | 7.4 | 0.328 | 0.959 | 99.04 |
| 8 | 85 | 8.7 | 0.616 | 1.610 | 98.39 |
| 9 | 85 | 8.3 | 0.483 | 1.160 | 98.84 |
| 10 | 85 | 6.6 | 0.277 | 0.876 | 99.12 |
| Control 1 | 85 | 7.4 | 1.030 | 1.270 | 98.73 |
| Control 2 | 85 | 7.6 | 0.251 | 0.544 | 99.46 |
| Control 3 | 85 | 7.3 | 0.356 | 0.954 | 99.05 |

**Table 6:** Halyard 62126 filter efficiency evaluation.

| Sample # | Flow Rate<br>(l min<sup>-1</sup>) | Initial Filter<br>Resistance<br>(mmH<sub>2</sub>O) | Initial<br>Leakage (%) | Max Leakage<br>(%) | Efficiency (%) |
| --- | --- | --- | --- | --- | --- |
| 1 | 85 | 10.8 | 1.880 | 1.920 | 98.08 |
| 2 | 85 | 10.7 | 1.660 | 1.730 | 98.27 |
| 3 | 85 | 10.1 | 1.660 | 1.670 | 98.33 |
| 4 | 85 | 10.5 | 1.170 | 1.320 | 98.68 |
| 5 | 85 | 10.8 | 2.510 | 2.690 | 97.31 |
| 6 | 85 | 10.7 | 1.340 | 1.370 | 98.63 |
| 7 | 85 | 10.5 | 2.090 | 2.140 | 97.86 |
| 8 | 85 | 10.9 | 1.681 | 0.681 | 99.32 |
| 9 | 85 | 10.5 | 1.660 | 1.660 | 98.34 |
| 10 | 85 | 10.5 | 0.768 | 0.768 | 99.23 |
| Control 1 | 85 | 11.7 | 1.040 | 1.060 | 98.94 |
| Control 2 | 85 | 11.5 | 0.348 | 0.352 | 99.65 |
| Control 3 | 85 | 11.4 | 0.968 | 0.971 | 99.03 |

#### 4.2.2 Manikin fit factor

Fit testing was run on 5 masks of each type, as well as 2 control masks. The respirator is mounted on a “static advanced headform,” which is connected to a breathing simulator. Two types of exercises are performed to simulate normal (11.2 l/min) and deep (20.4 l/min) breathing, during which fit factor is measured directly using a Portacount Pro+ model 8038 Respirator Fit Tester. Three successive exercises, in the sequence normal, deep, normal, are reported, and the overall fit factor is the harmonic mean of these tests. For more details see [5]. The results are summarized in Tables 7, 8, and 9.

**Table 7:** 3M 1860 manikin fit evaluation.

| Sample # | mFF Normal Breathing 1 | mFF Deep Breathing | mFF Normal Breathing 2 | Overall Manikin Fit Factor |
| --- | --- | --- | --- | --- |
| 11 | 200+ | 200+ | 200+ | 200+ |
| 12 | 200+ | 200+ | 200+ | 200+ |
| 13 | 200+ | 200+ | 200+ | 200+ |
| 14 | 200+ | 200+ | 200+ | 200+ |
| 15 | 200+ | 200+ | 200+ | 200+ |
| Control 4 | 200+ | 192 | 200+ | 198 |
| Control 5 | 200+ | 200+ | 200+ | 200+ |

**Table 8:** 3M 8511 manikin fit evaluation.

| Sample # | mFF Normal Breathing 1 | mFF Deep Breathing | mFF Normal Breathing 2 | Overall Manikin Fit Factor |
| --- | --- | --- | --- | --- |
| 11 | 200+ | 199 | 200+ | 200 |
| 12 | 200+ | 200+ | 200+ | 200+ |
| 13 | 200+ | 200+ | 200+ | 200+ |
| 14 | 200+ | 200+ | 200+ | 200+ |
| 15 | 200+ | 88 | 195 | 140 |
| Control 4 | 200+ | 200+ | 200+ | 200+ |
| Control 5 | 200+ | 200+ | 200+ | 200+ |

**Table 9:** Halyard 62126 manikin fit evaluation.

| Sample # | mFF Normal Breathing 1 | mFF Deep Breathing | mFF Normal Breathing 2 | Overall Manikin Fit Factor |
| --- | --- | --- | --- | --- |
| 11 | 189 | 150 | 200+ | 177 |
| 12 | 200+ | 200+ | 200+ | 200+ |
| 13 | 160 | 105 | 142 | 131 |
| 14 | 200+ | 148 | 197 | 178 |
| 15 | 128 | 80 | 180 | 116 |
| Control 4 | 200+ | 181 | 200+ | 194 |
| Control 5 | 120 | 159 | 85 | 114 |

#### 4.2.3 Strap integrity

The tensile strength testing was done on straps of 3 masks used for filtration testing, as well as the straps of 2 control masks. Top and bottom straps are tested separately. Straps are “pre-stretched” to twice their original length for 2 minutes, and released for 5 minutes, after which the new length is measured. Straps are then held at 150 % of their new length, and the force is reported after 30 s. More details on the test procedure can be found in [5], and the raw data for our tests are in Tables 10, 11, and 12.

**Table 10:** 3M 1860 strap integrity evaluation.

| Sample # | Force in top strap (N) | Force in bottom strap (N) |
| --- | --- | --- |
| 1 | 2.711 | 2.458 |
| 2 | 2.802 | 2.528 |
| 3 | 2.667 | 2.517 |
| Decontaminated Strap Average | 2.727 | 2.501 |
| Control 1 | 2.928 | 2.591 |
| Control 2 | 2.907 | 2.540 |
| Control Strap Average | 2.918 | 2.566 |

**Table 11:**
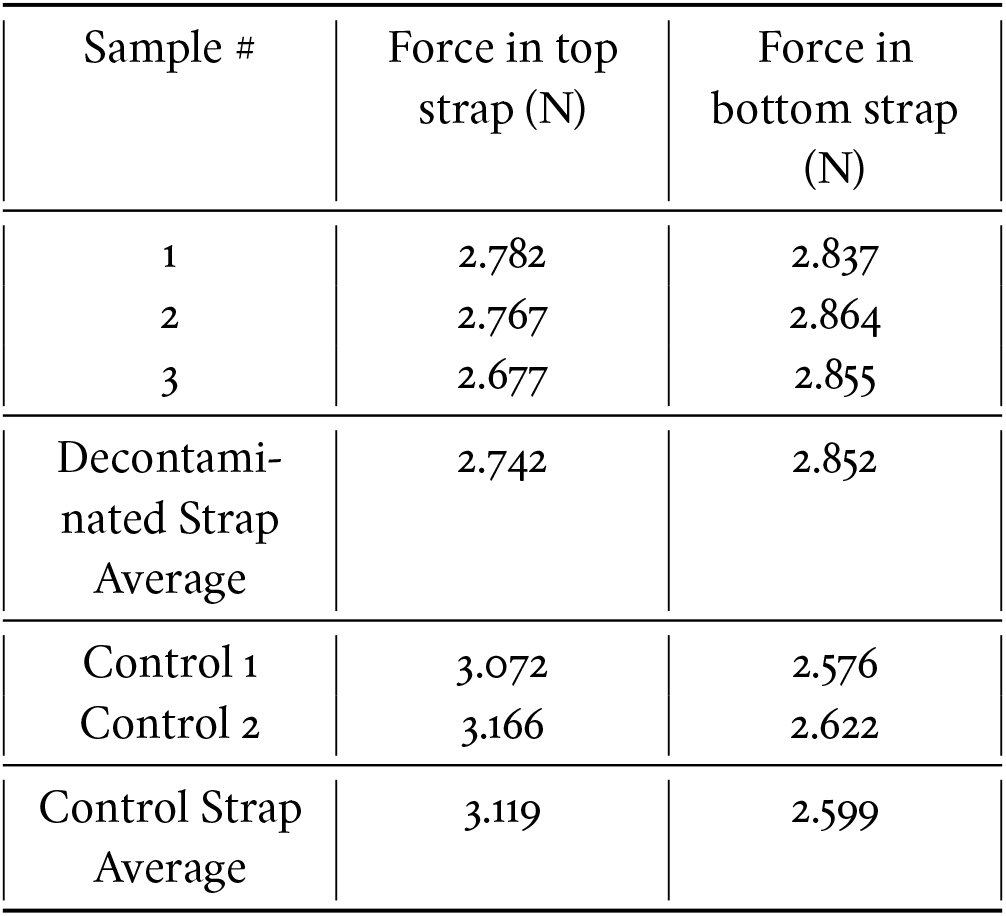
3M 8511 strap integrity evaluation.

**Table 12:** Halyard 62126 strap integrity evaluation.

| Sample # | Force in top strap (N) | Force in bottom strap (N) |
| --- | --- | --- |
| 1 | 2.144 | 2.187 |
| 2 | 2.132 | 2.209 |
| 3 | 2.181 | 2.268 |
| Decontaminated Strap Average | 2.152 | 2.221 |
| Control 1 | 2.283 | 2.343 |
| Control 2 | 2.228 | 2.345 |
| Control Strap Average | 2.256 | 2.344 |

As noted above, for the 3M 8511 mask, the bottom strap became stronger on average. We do not believe our decontamination method strengthens the straps, and attribute this effect to the relatively small sample size used for this test (3 treated masks and 2 controls).

### 4.3 Pathogen inactivation

To anchor our discussion of pathogen inactivation, we focus on the Food and Drug Administration (FDA) guidelines for Emergency Use Authorization (EUA) submission related to mask decontamination [8]. According to these guidelines, a decontamination method should result in at least a 3 log_10_ reduction of “viricidal activity.” Fischer et al. [1] used dry heat at 70 °C for 1 hour to heat a contaminated N95 FFR, resulting in *>* 3-log reduction of plaque forming units (PFUs) of SARS-CoV-2 deposited on the mask. This constitutes the most relevant current data point suggesting the inactivation potential of our method, which targets higher temperatures (at least 80 °C) and longer times (at least 65 min). (Higher temperatures and longer times are generally accepted to result in larger reductions of infective viral units.) Thus our method, even in the worst-case conditions, exceeds the requirements suggested by [1].

It is important to qualify this result with several caveats. First, a 3-log reduction of viral infectivity is the lower threshold of an acceptable decontamination procedure. Second, viral inactivation is highly sensitive to environmental conditions, such as the surface upon which the virus is deposited, the residual humidity, and the presence of other bodily fluids (such as saliva). We hope that further studies on heat inactivation with higher temperatures (such as our >80 °C target) will result in a further one to two-log reduction of the SARS-CoV-2 infectivity, which should help mitigate these concerns.

The FDA guidelines [8] at present also require 6-log reduction in either mycobactericidal or sporicidal activity. We have not performed a detailed evaluation of our method to check for appropriate reductions in bacterial or other contaminants; indeed we believe this method **will not** decontaminate certain other pathogens. For example, experiments [9] heating mycobacteria (including *M. tuberculosis*) for (15 to 30) min at 95 °C have shown up to 50 % survival. Another experiment [10] heats *M. tuberculosis* in culture for 20 min at 80 °C and finds less than 2-log reduction. It is possible that longer treatments at (85 to 95) °C will inactivate mycobacteria, but we have not found direct evidence to this effect in the existing literature. For this reason, we assume that this method only inactivates SARS-CoV-2 at the required 3-log level.

### 4.4 Availability of tested models

In this work, we only make statements particular to certain dryers manufactured by Alliance Laundry Systems (ALS). These dryers are particularly suited to the goals of our decontamination protocol, as ALS is the largest worldwide manufacturer of commercial laundry equipment and sells equipment to more than 140 countries. There are approximately 100 000 units in North America, and 47 000 internationally, that are compatible with this protocol (see Appendix A).

Other dryer models from other manufacturers may be equally suitable for a similar protocol. To evaluate a dryer, the temperatures achieved in decontamination cycles must be characterized as described in Section 4.1. If the equilibrium temperatures are suitably stable, consistent, and narrow, the settings of the machine should be calibrated so that the mask bag environment reaches temperatures of at least 80 °C for at least 60 min.

## 5 Outlook

The primary goal of this research is to develop an affordable, scalable, and reliable decontamination procedure for N95 FFRs using existing resources. By rigorously assessing a procedure tailored to certain widely available dryer models, the data-driven protocol outlined above passes the important tests of temperature stability and repeatability on a single machine. The data provided by the CDC/NIOSH FFR assessment task-force show minimal drop in filtration efficiency and satisfactory fit. This paves the way for an application with the FDA to obtain an Emergency Use Authorization.

While this article reports results for only two specific ALS dryer models, we hope that sharing our methodology and the many lessons learned during our experimental runs will empower other groups to perform similar testing on a wider range of models, including from other manufacturers.

## Data Availability

Raw data of temperature sensor logs used in this paper can be obtained by emailing. All other data is presented in the main text or appendices.

## 6 Acknowledgements

First, we especially thank Michael S. Bergman and his group at the NPPTL for providing key data for this work, without which no protocol could be published. We also thank Dale Greeson and his team, as well Joseph Zeman and Lara Simmons, at Medline for supplying the masks we used in this study. We would also like to highlight Jess Riedel who initially proposed using laundry dryers as a decontamination method. YL gives special thanks to Patricia Diaz and Omar Hernandez, as well as Shakeel Hakim, who provided access at the beginning of this project to industrial dryers. We thank Amita Gupta for feedback on the manuscript. YL, EM & DR would also like to thank Cheryne Jonay for collaboration at early stages of this project. YL is supported by the Hertz Foundation. EM would like to thank Sean Hartnoll at Stanford University for support and funding, and YL & DR likewise thank Xiao-Liang Qi at Stanford University.

## A Determining dryer model compatibility and settings

This decontamination method can only be used with models explicitly addressed in this appendix. Temperature settings for models whose decontamination cycle temperatures we have directly measured are listed in Table 3.

To tell if a particular dryer is compatible, first find the model identifier, as shown in Figure 7. The first six characters of this identifier must match one of the tumbler model numbers listed in Table 3. Next, not all dryers that have matching tumbler identifiers can be set to the temperatures and runtimes listed in Table 3. Only dryers with controls that allow run parameters matching those in the appropriate row of Table 3 can be used. This can either be checked by the user, or by checking the model identifier. If the model identifier has 15 characters, then characters 7 and 8 determine the control type, and should be one of EO, RE, UO, or RU. If the model identifier has 18 characters, only character 7 determines the control type, and should be D or N.

**Figure 7:**
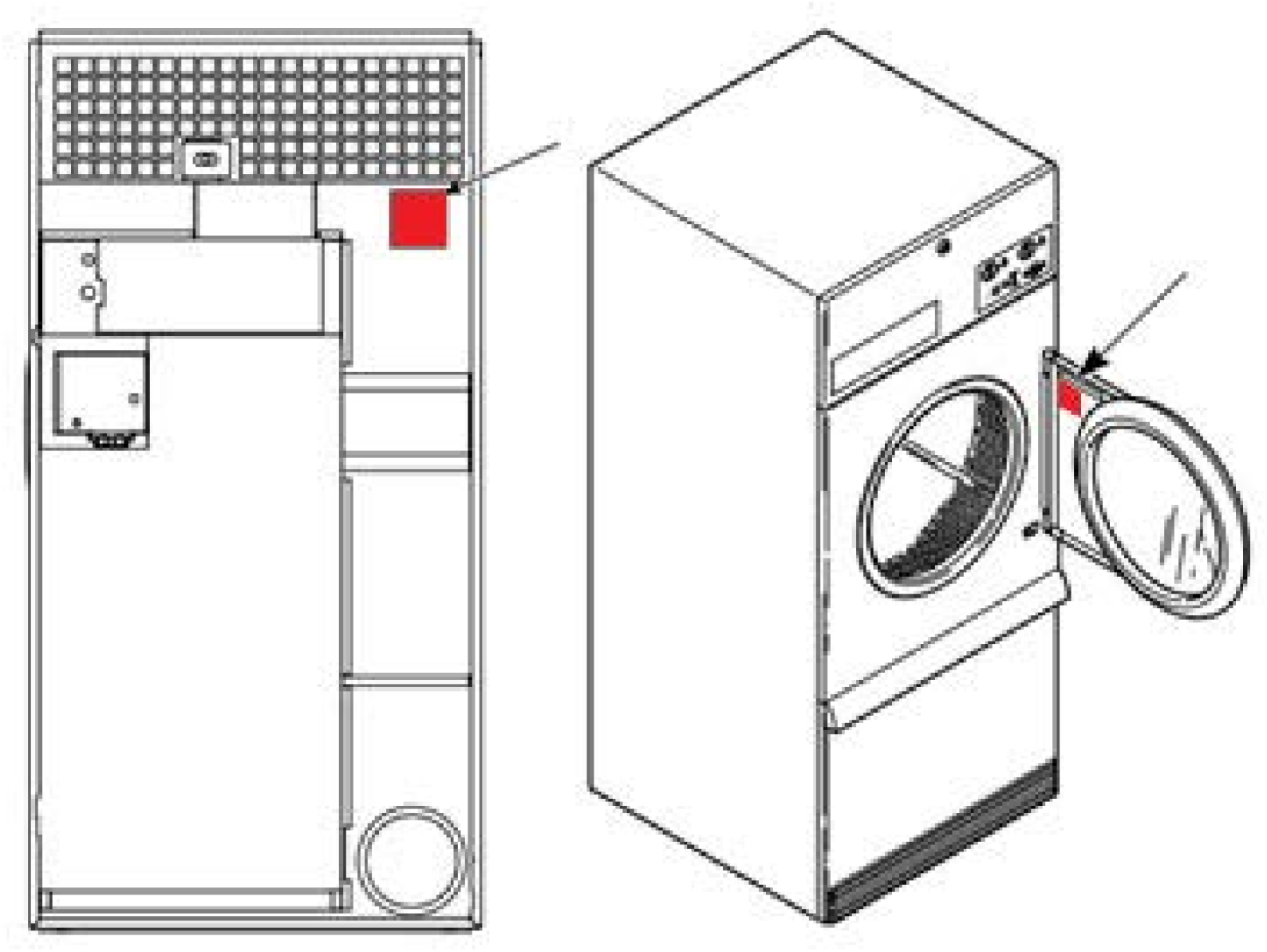
Model and serial numbers can be found when the loading door is opened on a sticker located on the inside of the hinge bracket. If that sticker is missing there is a serial plate on the back of the machine.

## B Raw data of NPPTL N95 FFR assessment

## Footnotes

1 Other bags that can be sealed may be effective, but should be tested without masks. Bags should not open during a decontamination cycle, and should remain airtight. Bags should not be reused over multiple decontamination cycles.

2 Inactivation of SARS-CoV-2 may be highly sensitive to the surface on which it resides. For instance, longer heating times may be needed to decontaminate metal surfaces than are needed for the surface of the mask [1]. Our heat protocol aims at decontamination of the filter piece of the mask. Disinfectant wipes should be used for the metallic nose piece present on some mask models. The EPA publishes and updates a list (“List N”) of disinfectant wipes recommended for use against SARS-CoV-2, located here as of July, 2020: https://cfpub.epa.gov/giwiz/disinfectants/index.cfm. We stress that we did not wipe the nosepieces of the masks sent to NPPTL for fit and filtration testing. While we expect this does not affect fit/filtration as long as the chemicals of the wipe do not come in contact with the N95 filtration material or the elastic straps, the data in this paper cannot be used to directly establish this.

3 These are over-estimates based on Plot A in the user manual.

4 In fact, the HOBO sensor we use is rated to come to equilibrium in 3.5 min in water; in air the equilibration time is much slower (10 min at 1 m s^−1^ airflow) so it is likely we are still overestimating our time to 80 °C.

5 The uncertainty in the mean efficiency is derived from the estimate of the standard deviation.

